# Trends and geographic variation in skin cancer incidence in Songkhla, Southern Thailand, 1989–2020: A Joinpoint regression and age-period-cohort analysis

**DOI:** 10.1101/2025.08.20.25334137

**Authors:** Suchaya Pajareeyaphan, Paramee Thongsuksai, Hutcha Sriplung, Wit Wichaidit

**Author notes:** **Corresponding author:** Suchaya Pajareeyaphan, MD, Faculty of medicine, Prince of Songkla University.

## Abstract

**Background:** The incidence trends of skin cancer are increasing across the world. However, data from Southeast Asian countries, including Thailand, are limited. Songkhla, a province in southern Thailand, has a predominant occupation and religion across different geographic areas. These differences may result in varying levels of UV exposure, potentially influencing the incidence of skin cancer. This study aimed to assess the trends in skin cancer incidence in Songkhla according to age, calendar period, birth cohort, and geographical area.

**Methods:** The study included patients who were incidently diagnosed with squamous cell carcinoma, basal cell carcinoma, and melanoma of the skin between 1989 and 2020, as recorded in the Songkhla population-based cancer registry. Geographic areas were classified into four categories according to predominant occupation and religion: farming and fishing area, city area, Muslim predominated area, and gardening area. Age-standardized incidence rates were calculated, and trend analyses were performed using Joinpoint regression and age-period-cohort (APC) analysis.

**Results:** The incidence of skin cancer in men remained unchanged before 2011; a similar pattern was observed in women before 2015. Thereafter, a significant decline was noted in both sexes, with annual percentage change rates of −9.53% in men and −11.51% in women. Based on geographic area, the highest incidence was observed in the city area (up to 7.14 in men, 4.62 in women), whereas the lowest was recorded in the Muslim predominated area (up to 3.93 in men, 3.32 in women). The APC analysis indicated that the incidence increased with age. The highest risk was observed in the cohort born in 1917, with a gradual decline in subsequent cohorts. No significant period effect was identified.

**Conclusions:** The incidence of skin cancer in Songkhla, Thailand, has declined over recent years. Variations in incidence are associated with differences in geographic region, occupation, and religious composition.

## 1. Introduction

Skin cancers can be divided into melanoma and nonmelanoma skin cancer (NMSC). The two primary subtypes of NMSC are basal cell carcinoma (BCC) and squamous cell carcinoma (SCC). According to GLOBOCAN 2022, non-melanoma skin cancer (excluding BCC) is the third most common cancer in men and the seventh in women, with age-adjusted incidence rates (ASRs) of 14.0 and 7.5 per 100,000, respectively. By contrast, melanoma is less prevalent, with ASRs of 3.7 and 2.9 per 100,000 in men and women, respectively (1). The incidence of NMSC varies significantly across countries, with the highest reported in Australia (2). In Thailand, the ASRs for melanoma and NMSC in 2016–2018 were 4.3 in women and 4.0 in men, whereas higher rates were observed in Songkhla province (4.9 in women and 4.8 in men) (3). A rising trend in the incidence of skin cancer has been reported in various regions worldwide, including Australia, European countries, and Hong Kong (4–6). However, data on incidence trends in Southeast Asian countries, including Thailand, remain limited.

Ultraviolet (UV) exposure is a well-known risk factor for skin cancer (7–9). An increasing UV index over time, due to stratospheric ozone depletion, has been correlated with the rising incidence of skin cancer in Australia (10). Individuals engaged in outdoor occupations, such as farming and fishing, are more likely to experience higher levels of UV exposure. Additionally, clothing style may influence the degree of UV exposure. However, no data have demonstrated whether these factors directly affect the incidence of skin cancer.

Songkhla Province, located in southern Thailand, comprises 16 districts with predominant occupations varying by geographical area, such as farming, fishing, gardening, and salaried employment. The religious composition also differs across regions, with both Buddhist and Muslim populations represented. These demographic and occupational variations may result in differing levels of UV exposure across districts, potentially influencing the incidence of skin cancer. Aging is another recognized factor associated with the development of skin cancer (7–9). In Hong Kong, population aging has been identified as a contributor to the increasing trend in skin cancer incidence (6). Thailand is similarly undergoing a demographic shift toward an aging society (11). Therefore, population aging may also play a role in the observed trends in skin cancer incidence.

Describing the trends in skin cancer incidence based on the geographic areas and potential risk factors may provide valuable insights for healthcare planning aimed at reducing and preventing skin cancer. Thus, the present study aimed to assess trends in skin cancer incidence in Songkhla, Thailand, by age, calendar period, birth cohort, and geographic area.

## 2. Materials and Methods

### 2.1. Data source

Patient data from the Songkhla population-based cancer registry for the period 1989–2020 were obtained from the Division of Digital Innovation and Data Analytics, Faculty of Medicine, Prince of Songkla University. The data were accessed on 21 October 2024. The data that could identify the patients were not assessed. We included the records of patients diagnosed with skin cancer using the International Classification of Diseases, 10^th^ revision, codes: C43 for malignant melanoma of the skin and C44 for other malignant neoplasms of the skin. For cases coded as C44, only SCC and BCC were included, identified using the International Classification of Diseases for Oncology, 3^rd^ edition, codes 805–808 for squamous cell neoplasms and 809 for basal cell carcinoma. Data including age, sex, date of birth, date of diagnosis, religion (Buddhist, Muslim, and others), residential district, histologic type, tumor site (face, body, and acral area), and disease stage (local/regional/distance) were collected, all of which were recorded in cancer registry forms. Patients with no data on date of diagnosis, age, and sex were excluded, as the incidence rates could not be calculated. This study was approved by the ethics committee of the Faculty of Medicine, Prince of Songkla University.

### 2.2. Statistical analysis

#### 2.2.1. ASR

The age-specific incidence rate for each year (1989–2020) was calculated based on 18 distinct age groups (categorized in 5-year intervals, ranging from 0–4 to ≥85 years old). These rates were subsequently adjusted to the world standard population to calculate the ASRs.

The population denominators for Songkhla were obtained from the Department of Public Administration, the Report of the Population Projections for Thailand 2010–2040 by the Office of the National Economic and Social Development Council (12), and the database of the Department of Provincial Administration (13).

The ASRs for all patients with skin cancer were calculated for each subgroup by sex and geographic area. The 16 districts of Songkhla were grouped into farming and fishing areas (Ranot, Krasae Sin, Sathing Phra, and Singhanakhon), city areas (Mueang Songkhla and Hat Yai), Muslim predominated areas (Chana, Thepha, and Sabayoi), and gardening areas (Khuan Niang, Rattaphum, Bangklam, Na Mom, Khlong Hoi Khong, Sadao, and Na Thawi), as illustrated in Figure 1.

**Figure 1.**
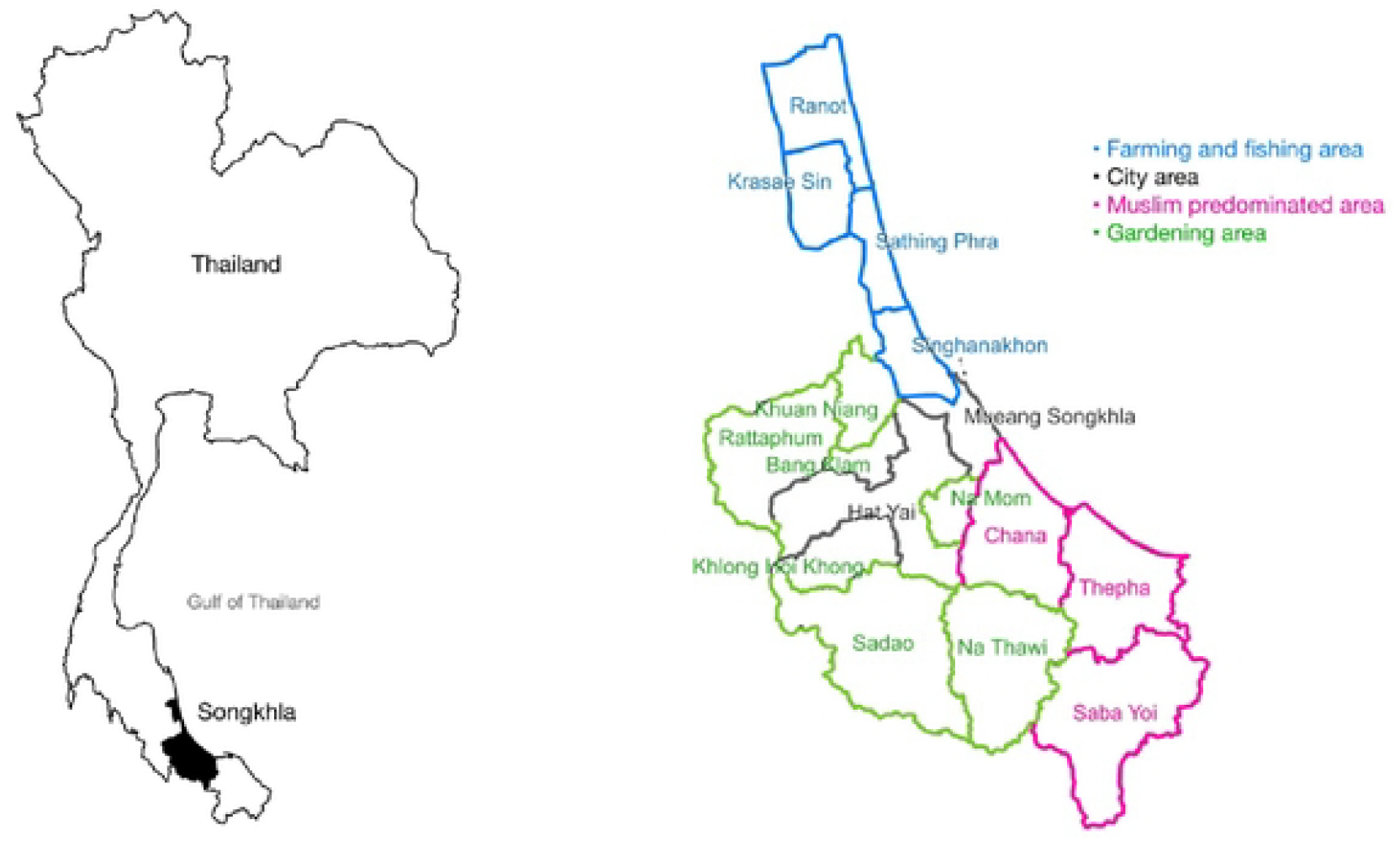
Map of Songkhla provinces showing all 16 districts

### 2.2.2. Trend analysis

Trends in skin cancer incidence were assessed using Joinpoint regression analysis (Joinpoint Regression Program, version 5.1.0; National Cancer Institute, Bethesda, MD, USA) to identify significant changes in trend, referred to as joinpoints. The effects of age, calendar year of diagnosis, and birth cohort on cancer incidence were examined using an age-period-cohort (APC) regression model.

## 3. Results

### 3.1 Demographic and clinical characteristics and overall annual incidence of skin cancer

A total of 1,651 patients were diagnosed with skin cancer, including melanoma, squamous cell carcinoma, and basal cell carcinoma, between 1989 and 2020. The characteristics of patients with incident cases are presented in Table 1. The number of men and women was nearly equal. The median age at diagnosis was 69 years. The most common cancer types were squamous cell carcinoma in men and basal cell carcinoma in women. The head and neck were the most common cancer sites.

**Table 1.** Demographic and clinical characteristics of male and female patients with skin cancer in Songkhla, 1989–2020.

| Characteristics | Number (%) |
| --- | --- |

|  | Men<br>(n = 822) | Women<br>(n = 829) |
| --- | --- | --- |
| Age (years) (median ± IQR) | 69 ± 19 | 69 ± 20 |
| **Religion** |  |  |
| Buddhist | 701 (85.3) | 738 (89.0) |
| Muslim | 102 (12.4) | 81 (9.8) |
| Christian | 15 (1.8) | 8 (1) |
| Other | 2 (0.2) | 2 (0.2) |
| Unknown | 2 (0.2) |  |
| **Address** |  |  |
| Hatyai | 244 (29.7) | 252 (30.4) |
| Mueangsongkhla | 146 (17.8) | 147 (17.7) |
| Ranot | 67 (8.2) | 50 (6.0) |
| Sadao | 57 (6.9) | 51 (6.2) |
| Singhanakhon | 56 (6.8) | 58 (7.0) |
| Chana | 48 (5.8) | 56 (6.8) |
| Sathingphra | 47 (5.7) | 42 (5.1) |
| Rattaphum | 32 (3.9) | 34 (4.1) |
| Nathawi | 27 (3.3) | 23 (2.8) |
| Bangklam | 22 (2.7) | 22 (2.7) |
| Thepha | 22 (2.7) | 25 (3.0) |
| Khuanniang | 14 (1.7) | 26 (3.1) |
| Sabayoi | 14 (1.7) | 11 (1.3) |
| Khlonghoikhong | 11 (1.3) | 10 (1.2) |
| Krasaesin | 8 (1.0) | 10 (1.2) |
| Namom | 7 (0.9) | 12 (1.4) |

| Cancer location |  |  |
| --- | --- | --- |
| head and neck | 327 (39.8) | 486 (58.6) |
| Lower limb and hip | 211 (25.7) | 130 (15.7) |
| Trunk | 138 (16.8) | 96 (11.6) |
| Upper limb and shoulder | 103 (12.5) | 68 (8.2) |
| Overlapping | 3 (0.3) | 6 (0.7) |
| Not otherwise specified | 40 (4.9) | 43 (5.2) |
| Skin cancer type |  |  |
| Squamous cell carcinoma | 448 (54.5) | 307 (37.0) |
| Basal cell carcinoma | 227 (33.7) | 434 (52.4) |
| Melanoma | 97 (11.8) | 88 (10.6) |

The incidence of skin cancer between 1989 and 2020 is shown in Figure 2, with ASRs ranging from 1.77 to 5.66 in men and from 1.87 to 5.52 in women. The ASR was higher in men than in women in almost all years, except for 1989, 1992, and 1996. The highest ASR was recorded in 1996 for women and in 2001 for men.

**Figure 2.**
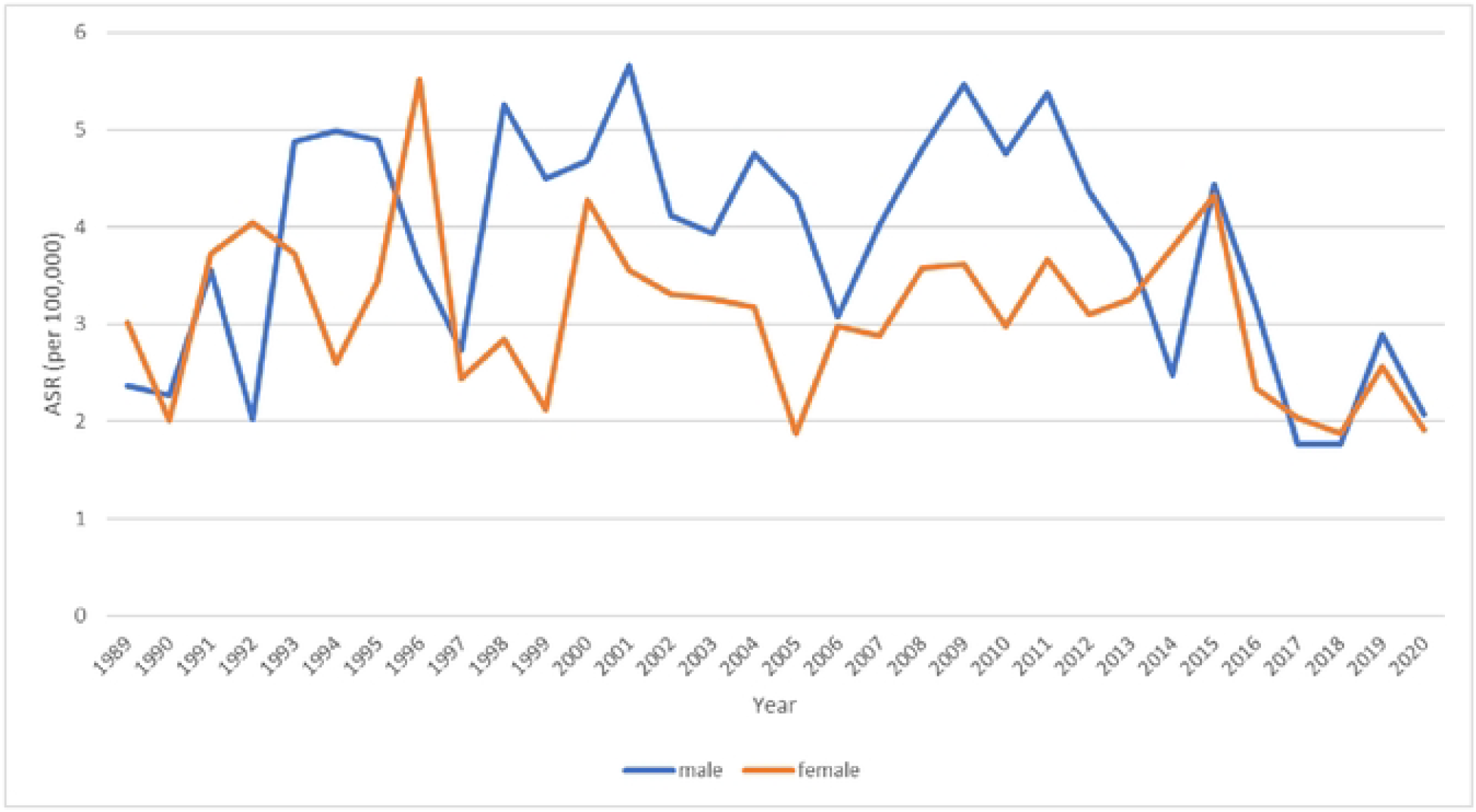
Age-standardized incidence rate (ASR) of first diagnosed skin cancer per 100,000 population by sex in Songkhla, Thailand, from 1989 to 2020

### 3.2. Incidence trend by sex and geographic area

From 1989 to 2011, the ASR of skin cancer among men showed an increasing trend before declining sharply (Figure 3). In 1989, the ASR in men was 3.62 per 100,000 population, rising to 4.96 in 2011, with an annual percentage change (APC) of 1.45% (p=0.067). After 2011, the ASR decreased to 2.01, with an APC of −9.53% (p<0.001). Among women, the ASR remained relatively stable from 1989 to 2015 (3.32– 3.36), with an APC of 0.04% (p=0.669). However, from 2015 onward, the ASR declined to 1.82 in 2020, with an APC of −11.51% (p=0.005).

**Figure 3.**
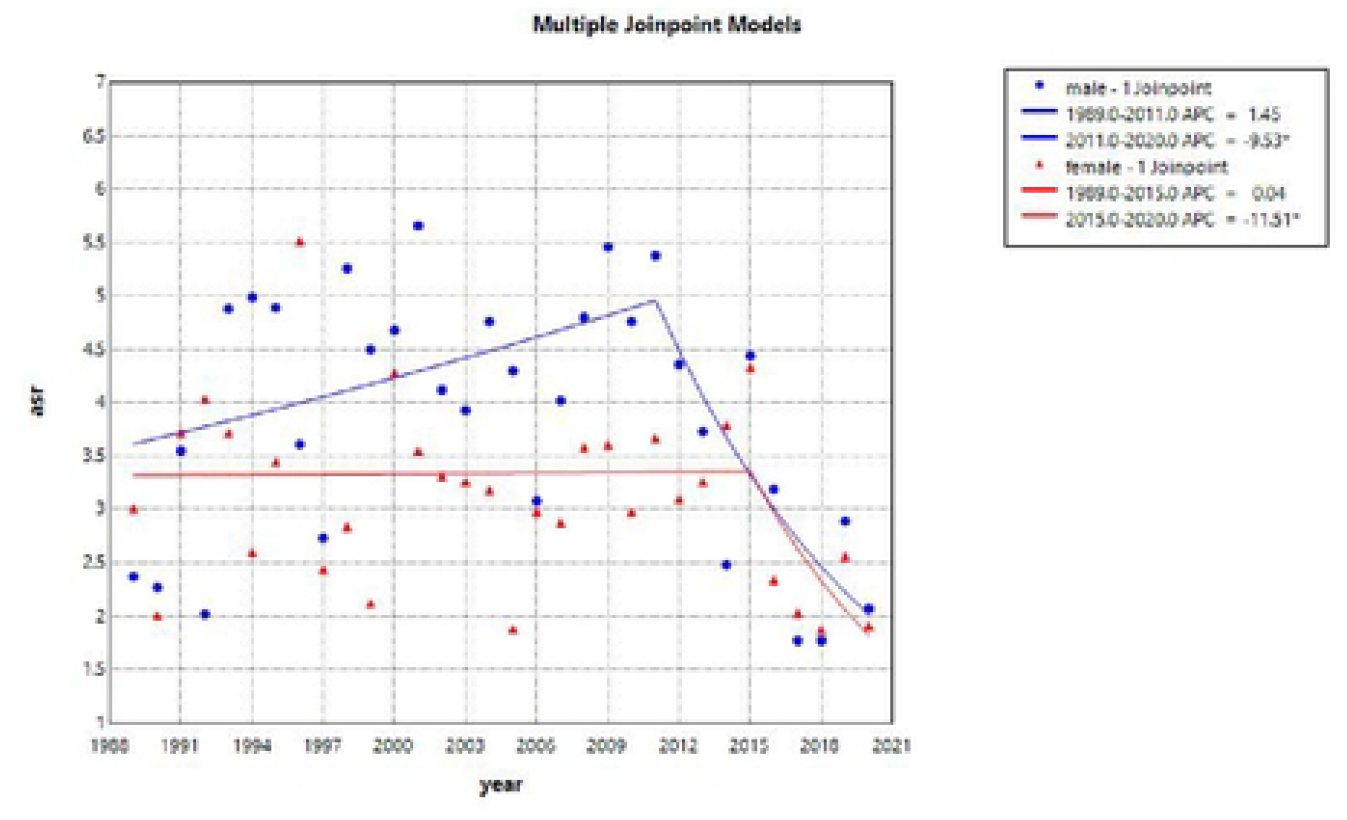
Trends in skin cancer incidence by sex in Songkhla, Thailand, from 1989 to 2020, based on the Joinpoint regression analysis

The incidence trends by sex and geographic area are illustrated in Figure 4. Among the male population, the highest ASR was observed in the city area (3.10–7.14), followed by the fishing and farming area (3.67–6.42), the gardening area (1.18–4.90), and the Muslim predominated area (2.24– 3.93). Slight decreasing trends were observed in the fishing and farming area and the Muslim predominated area, with APC rates of −2.40 (p=0.170) and −2.43 (p = 0.080), respectively. By contrast, the trends in the city and gardening areas slightly increased before 2011, with APC rates of 0.55 (p=0.576) and 1.31 (p=0.503), respectively. Following 2011, both areas experienced significant declines, with APC rates of −8.86 (p=0.03) and −14.61 (p=0.012), respectively.

**Figure 4.**
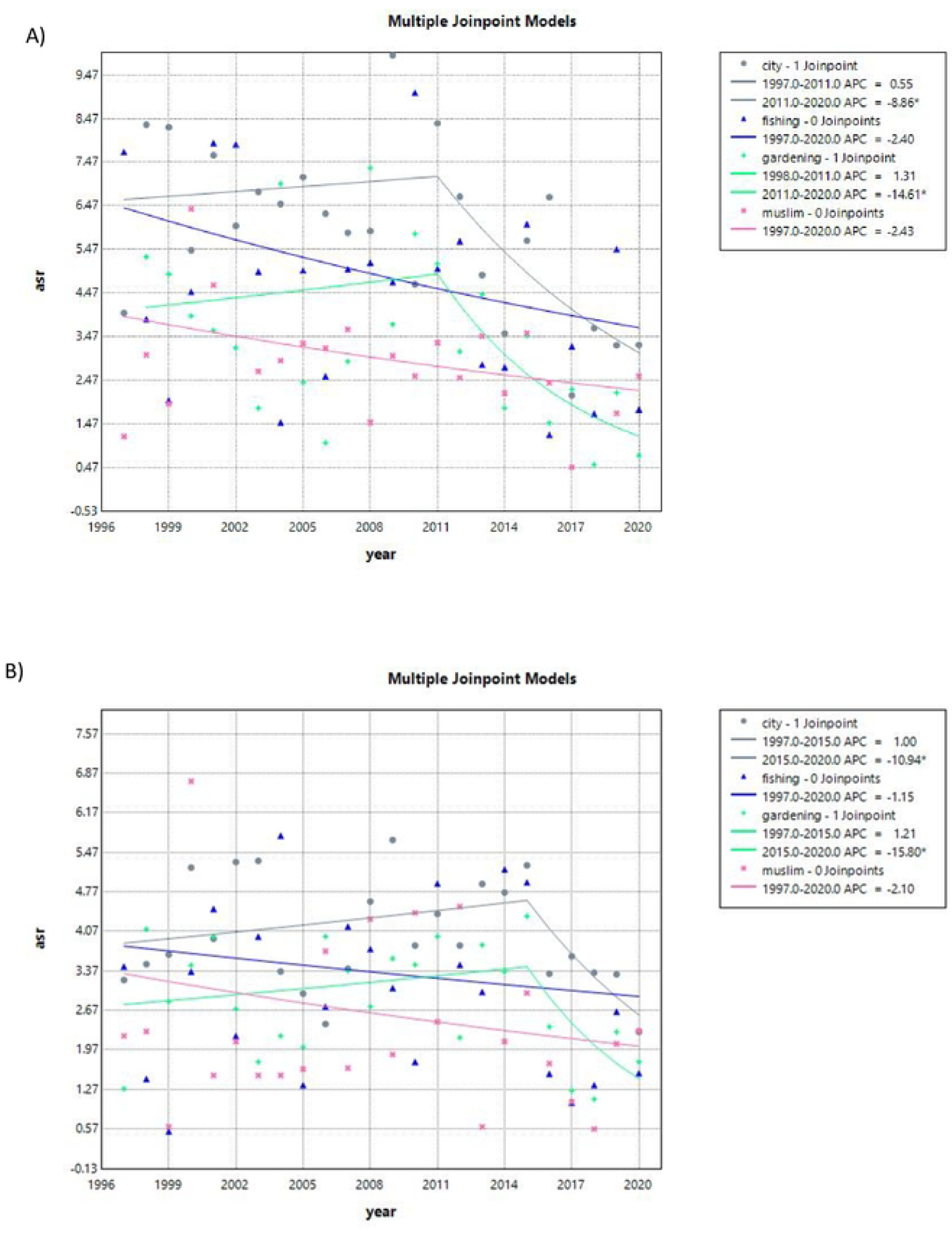
Trends in skin cancer incidence by geographic area in Songkhla, Thailand, from 1989 to 2020 based on the Joinpoint regression analysis. A) Male population. B) Female population

In the same way, for the female population, the city area had the highest ASR (2.59–4.62), followed by the fishing and farming area (2.92–3.81), the gardening area (1.46–3.45), and the Muslim predominated area (2.04–3.32). Incidence trends in the fishing area and the Muslim predominated area slightly decreased, with APC rates of −1.15 (p=0.594) and −2.10 (p=0.307), respectively. Meanwhile, the trends in the city and gardening area slightly increased before 2015, with APC rates of 1.00 (p=0.186) and 1.21 (p=0.261), respectively. After 2015, the trends in both areas significantly decreased, with APC rates of −10.94 (p=0.016) and −15.80 (p=0.030).

### 3.3. Age-period-cohort analysis

The results of the APC, age-cohort (AC-P), and age-period (AP-C) models were similar for both men and women (Figure 5). The APC model showed that the incidence of skin cancer increased with age. With regard to the cohort effect, individuals born in 1917 exhibited the highest risk, with a gradual decline in risk observed among younger cohorts. In contrast, no significant changes in risk were associated with calendar year.

**Figure 5.**
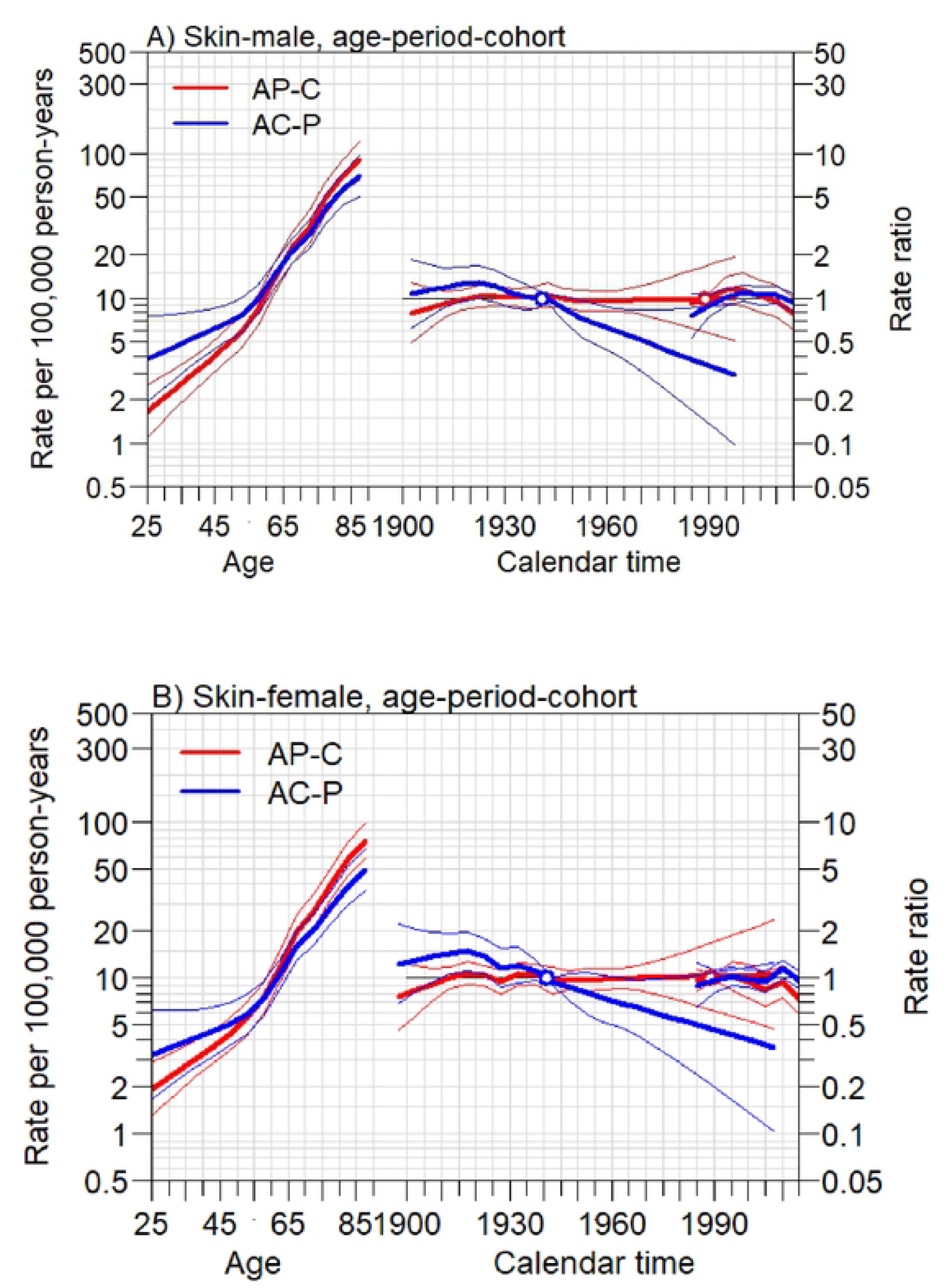
Trends in skin cancer incidence in Songkhla, Thailand, from 1989 to 2020 based on the age-period-cohort model (left), age-cohort model (AC-P, middle), and age-period model (AP-C, right). A) male population. B) Female population

## 4. Discussion

The incidence of skin cancer in men remained relatively unchanged prior to 2011 and in women prior to 2015. Subsequently, the incidence trends decreased significantly in both sexes. Based on geographic area, the highest incidence was recorded in the city area, whereas the lowest was observed in the Muslim predominated areas. The APC analysis demonstrated that the incidence rate increased with age. The cohort born in 1917 exhibited the highest risk ratio, which gradually decreased in the younger cohorts. The period effect did not have a significant impact on the risk ratio.

The incidence of skin cancer was slightly higher in men than in women (1.44–5.38 vs 1.77–4.77), which is consistent with the patterns reported in other countries (2). The incidence trend in men remained relatively stable until 2011 and in women until 2015. Subsequently, a significant decline in the incidence of skin cancer was observed in both sexes. This pattern contrasts with trends reported in many countries, such as Western countries, Australia, and Hong Kong, where the incidence has gradually increased (4–6). In the United States, the incidence trends have remained relatively stable (14). In Western countries, the increasing incidence has been attributed to the increased participation in artificial tanning and holidays in sunny climates, leading to increased exposure to UV radiation. In some countries with a high incidence of skin cancer, such as Australia, public health campaigns have been implemented to raise awareness, promote self-examination, and encourage seeking medical attention. These efforts have contributed to increased skin cancer detection. However, according to culture and traditions in Thailand, people prefer fair skin; therefore, they usually avoid sun exposure, and campaigns related to skin cancer are quite limited. This may be one reason why the incidence of skin cancer in Thailand has not increased.

The incidence trends in Songkhla, Thailand, significantly decreased after 2011 in males and in 2015 in females. This could be due to the effects of changes in UV radiation that decreased after the 2000s, as UV radiation was the major risk factor for skin cancer. According to Bais et al., who studied UV radiation changes with the impact of ozone and clouds, ozone depletion has stabilized since the late 1990s and has started to recover. UV radiation increased from the 1960s, reached a broad maximum during the 2000s, and gradually decreased thereafter(15).

The incidence of skin cancer by geographic area revealed the highest rate in the city area, followed by the fishing and farming area, gardening area, and Muslim predominated area, respectively. These variations suggest that the occupational environment and clothing practices may have affected the incidence. Muslim individuals, particularly women, typically wear clothing that covers most of the body, resulting in reduced exposure to UV radiation. The gardening area in Songkhla primarily consists of rubber plantations, which provide shade during working and may reduce exposure to UV radiation. By contrast, individuals residing in fishing and farming areas—particularly those working in rice fields—are generally exposed to greater amounts of sunlight, resulting in a higher incidence of skin cancer. The highest incidence was observed in the city area. This may be attributed to multiple factors. Urban populations are generally more highly educated. Moreover, the presence of three large tertiary hospitals in the city area facilitates better access to healthcare services.

According to the APC analysis, the incidence rate of skin cancer increased with age. The cohort effect peaked in individuals born around 1917 and then steadily declined in successive younger cohorts. Meanwhile, the period effect demonstrated no significant impact on the rate ratio. The age effect was comparable, but the cohort and period effects differed from those reported in a study conducted in Hong Kong—the only study to assess skin care incidence trends using APC analysis (6). In that study, the cohort effect peaked in the 1975 birth cohort, and the period effect showed a continuous increase in the rate ratio over time. These trends were attributed to heightened public and physician awareness of skin cancer. However, the highest rate ratio in Songkhla was observed in the 1917 birth cohort. This may be explained by the fact that individuals born in 1917 reached older, cancer-susceptible ages around 1987, a period that coincided with increasing UV radiation levels, according to Bais et al. (15). Moreover, before the 2000s, agriculture—an occupation associated with considerable exposure to UV radiation— was the most prevalent form of employment in Thailand. During this period, the use of sunscreen was not widespread. However, the rate ratio did not change significantly across calendar periods. This may be attributed to the absence of major national campaigns or policies focused on skin cancer prevention in Thailand.

## 5. Conclusion

In conclusion, a decreasing trend in the incidence of skin cancer has been observed in Songkhla. This decline may be influenced by changes in UV radiation levels, occupational patterns, and cultural and traditional practices. Furthermore, variations in geographical areas—characterized by differing occupations and religious practices—appear to impact the incidence of skin cancer.

## Data Availability

All relevant data are within the manuscript and its Supporting Information files.

## Funding

### Declarations of interest

### Author contributions

### Data availability statement

## Abbreviations

NMSC: nonmelanoma skin cancer
BCC: basal cell carcinoma
SCC: squamous cell carcinoma
ASR: age-standardized incidence rate
UV: Ultraviolet
APC: age-period-cohort

## Notes

### Competing Interest Statement

The authors have declared no competing interest.

### Funding Statement

The author(s) received no specific funding for this work.

### Author Declarations

This study was approved by the ethics committee of the Faculty of Medicine, Prince of Songkla University. (REC.66-523-5-1)

